# High-dose accelerated intermittent theta burst stimulation improves cognitive function in early Alzheimer’s disease: A randomized sham-controlled trial

**DOI:** 10.64898/2026.02.13.26346250

**Authors:** Na Xu, Yi Xing, Aonan Li, Ruiqi Pan, Shuaicheng Liu, Jing Gao, Xinyu Liu, Tingyun Tao, Ping Zhang, Wuxiang Xie, Ning Guo, Yue Chen, Xiaoyue Sun, Jilin Wu, Weijun Gong, Hesheng Liu, Yi Tang, Danhong Wang

## Abstract

**Introduction:** This clinical trial investigates the efficacy and safety of a personalized 15-day accelerated intermittent theta-burst stimulation (aiTBS) protocol, targeted at either the default mode network (DMN) or the fronto-parietal network (FPN), in individuals with mild Alzheimer’s disease (AD).

**Methods:** 45 patients with mild AD were randomized 1:1:1 to receive 15 consecutive days of high-dose aiTBS (7200 pulses/day) targeting the DMN or FPN, or sham. The primary outcome was the change in ADAS-Cog after 15 days of treatment.

**Results:** Both active aiTBS groups demonstrated significantly greater ADAS-Cog improvement than sham at the primary endpoint. Response rates for a clinically meaningful improvement (≥3-points on ADAS-Cog) were significantly higher in the active groups (DMN: 38%; FPN: 47%) than in the sham group (0%). The improvement in active groups was sustained at 3-month follow-up.

**Discussion:** Personalized aiTBS targeting the DMN or FPN produced clinically meaningful cognitive benefits in mild AD and was safe.

## 1. Background

Alzheimer’s disease (AD) is characterized by large-scale brain network dysfunction^1^, with the default mode network (DMN) and fronto-parietal network (FPN) representing epicenters where the convergence of pathology, atrophy, and disconnection drives cognitive decline.^2–6^ The disease process triggers a cascading dysfunction across these networks, beginning typically in posterior DMN regions and progressively invading prefrontal regions of both the DMN and FPN.^7,8^ Within this architecture, the prefrontal nodes of the DMN and FPN are particularly important for episodic retrieval, executive control, working memory, and the higher-order integration of cognitive operations.^9,10^ Given their pivotal role, these hubs represent promising targets for circuit-based AD therapies.

Repetitive transcranial magnetic stimulation (rTMS), a non-invasive neuromodulatory approach, has emerged as a promising therapy due to its favourable safety profile^11–13^ and its potential in improving cognitive functions in AD.^14–17^ However, the generalization of its application is hampered by significant methodological heterogeneity, particularly in target precision and stimulation dosage, as repeatedly noted in meta-analyses^18–20^ and guidelines.^12^ While many rTMS trials aim to engage nodes of the DMN or FPN, few employ network-defined, individualized targets.^15^ The efficacy of neuromodulation depends critically on targeting functionally relevant neural circuits.^21,22^ However, both the FPN and DMN exhibit substantial inter-individual variability in functional organization^23,24^, thus standardized anatomical targeting may fail to recruit the intended network.^25,26^ The lack of target precision likely contributes to the variable responses reported in TMS studies that stimulate broad regions like the dorsal lateral prefrontal cortex (DLPFC).^16,27,28^ To address this gap, we employed the precision functional mapping approach^23^ to define patient-specific DMN targets in the dorsal medial prefrontal cortex (DMPFC) and FPN targets in the DLPFC. This allowed us to directly target circuit-level dysfunction and test its therapeutic potential for AD.

Conventional rTMS protocols for cognitive enhancement in AD typically follow a once-daily treatment spanning 2–6 weeks, with meta-analytic evidence indicating that ≥3 weeks and ≥20 sessions yield the most significant cognitive benefits.^20^ Yet long courses pose practical barriers for older patients. Accelerated paradigms using intermittent theta-burst stimulation (iTBS)^29^, which condenses multiple daily sessions into a shorter overall timeframe, have emerged as a promising alternative. The clinical potential of this approach is supported by its success in major depressive disorder (MDD), where an intensive 5-day regimen (10 daily sessions, 18000 pulses/day) has received FDA approval due to its rapid and robust antidepressant efficacy.^30^ In AD, two preliminary studies^31,32^ applying aiTBS to the DLPFC have reported encouraging cognitive results at daily doses of 1800 and 3600 pulses, respectively, although DMN targets remain unexplored using this paradigm.

In the present study, we conducted a randomized, sham-controlled, three-arm trial to test whether a 15-day, personalized, aiTBS protocol (4 daily sessions, 7200 pulses/day) —targeting the prefrontal DMN node within the dorsomedial prefrontal cortex (DMPFC) or the FPN node in the DLPFC—would improve cognition in mild AD.

## 2. Methods

### 2.1. Study Design

This study was a monocentric, randomized, sham-controlled, parallel-group clinical trial investigating the efficacy and safety of aiTBS in mild AD. Eligible participants were randomly assigned to one of three intervention arms: active aiTBS to the DMN, active aiTBS to the FPN, or sham stimulation. Treatment spanned 15 consecutive days and comprised four sessions per day (two in the morning and two in the afternoon). The study was registered at ClinicalTrials.gov (NCT05872243) in May 2023 and conducted in full compliance with the Declaration of Helsinki.

### 2.2. Participants

Patients were recruited from the Department of Neurology of the Xuanwu Hospital, Capital Medical University, China. Inclusion criteria were: (1) probable Alzheimer disease according to National Institute on Aging–Alzheimer’s Association criteria with amyloid positivity on positron emission tomography (PET) or cerebrospinal fluid (CSF); (2) Mini-Mental State Examination (MMSE) score 20-26; (3) Clinical Dementia Rating (CDR) score of 1; (4) age 50-90 years; (5) stable Alzheimer’s disease medication for at least 3 months; (6) availability of a reliable caregiver; and (7) ability to understand study procedures and provide written informed consent (or consent by a legal representative). Exclusion criteria included: (1) non-AD causes of cognitive impairment; (2) significant focal brain lesions on MRI; (3) significant head trauma or other major neurological disorders; (4) alcohol or substance abuse; (5) personal or first-degree family history of seizures; (6) any TMS or MRI contraindications. All participants or their legal guardians provided written informed consent.

### 2.3. Randomization and Blinding

Participants were randomized in a 1:1:1 ratio to receive active aiTBS to the DMN or FPN, or sham stimulation using stratified design to minimize imbalance in age (<75 vs. ≥ 75 years) and education level (<12 vs. ≥ 12 years). An independent statistician (WX) generated the allocation sequence using computer-generated random codes. The assignments were concealed using sequentially numbered, opaque, sealed envelopes, which were prepared and managed by a research assistant not involved in the trial. Sham stimulation used a coil identical in appearance and acoustic characteristics to the active coil. The TMS operator had no role in study design, outcome assessment, or data analysis and interacted with participants only regarding comfort or adverse effects. Participants, outcome assessors, therapists, data analysts, and other research staff were blinded to group assignment throughout the trial.

### 2.4. Procedures

Participants received four iTBS (or sham) sessions per day (7,200 pulses per day)—two in the morning and two in the afternoon—with a 50-minute intersession interval within each half-day, for 15 consecutive days. Stimulation intensity was 100% of the resting motor threshold (RMT). Treatments were delivered with neuronavigated TMS stimulators (MT20A, Neural Galaxy Inc, Beijing) equipped with double-sided coils. All patients continued on a stable background of standard drug therapy, which was not altered throughout the trial.

### 2.5. MRI Data Acquisition and Preprocessing

High-resolution T1-weighted structural images and resting-state functional MRI were acquired on a Siemens Spectra 3T MRI scanner. We collected 30 minutes of fMRI data per participant to ensure reliable subject-specific functional network parcellation. Imaging data were preprocessed using pBFS Cloud (Neural Galaxy, Beijing) according to the procedure previously described.^33^

### 2.6. Personalized Parcellation and Target Localization

The personalized targeting methodology for the DMN and FPN networks is illustrated in Figure 1. Following preprocessing of the MRI data, the cerebral cortex was first parcellated into fine-grained functional regions.^23^ The DMN target was searched in the left DMPFC, and the FPN target was searched in the left DLPFC. The final stimulation target within each search area was selected through a dual-criterion approach that balanced functional relevance and practical feasibility. Functional relevance was quantified by the degree of similarity between a voxel’s functional connectivity profile and the topology of the target network (DMN or FPN) derived from the parcellation, while feasibility was determined by sulcal depth to ensure the site was accessible for TMS.

**Figure 1.**
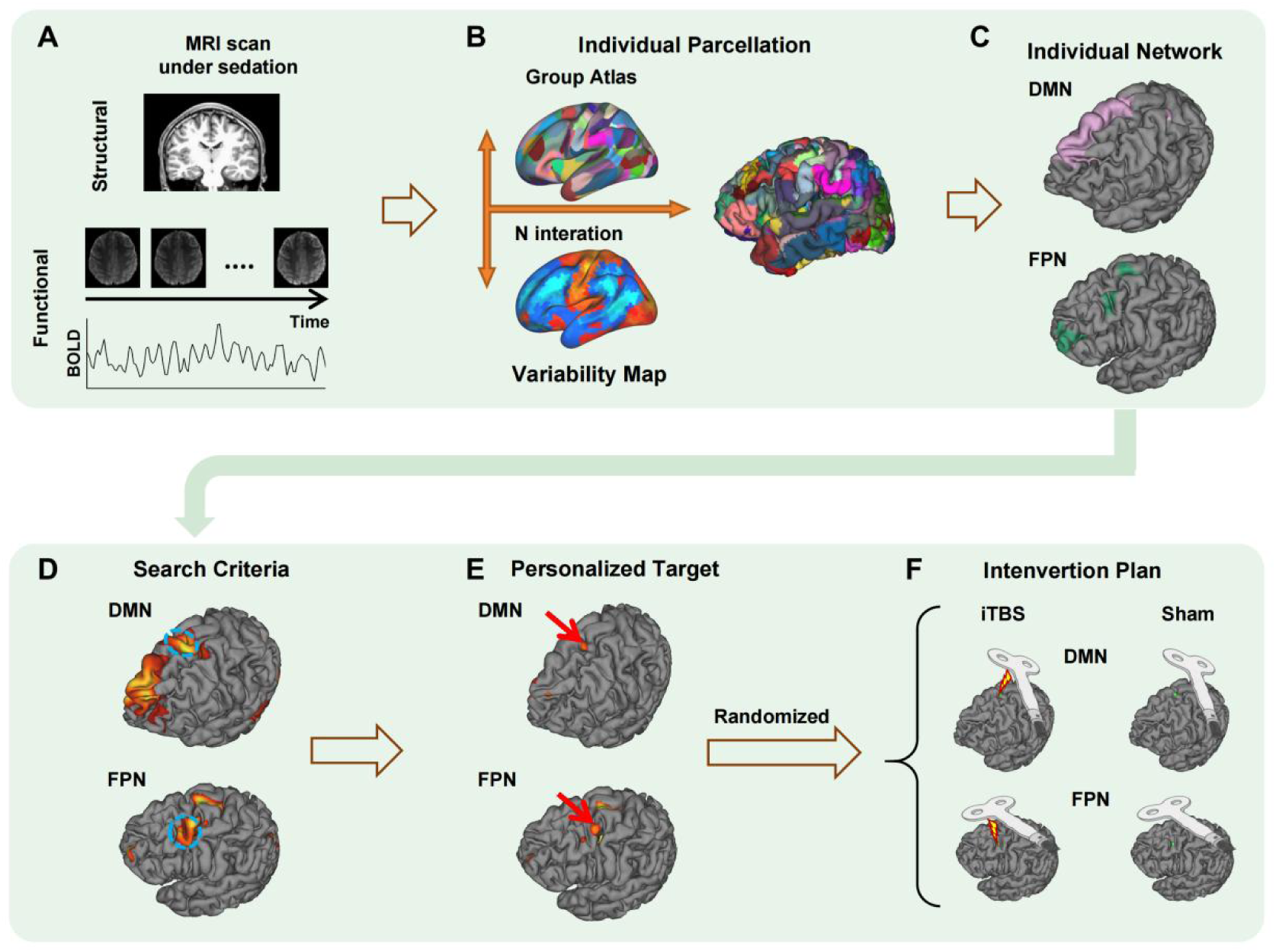
Trial procedure. Personalized DMN or FPN target identification. (A) For each participant, baseline structural and 30-min resting-state functional MRI data were acquired. (B) Individual functional parcellation was generated using an iterative approach previously reported, which leverages both the group-level atlas and an inter-individual variability map to produce vertex-wise confidence estimates for network assignment. (C) Frontal portions of the DMN and FPN were identified within the DMPFC and DLPFC, respectively. (D) Candidate stimulation sites were selected by weighing network functional connectivity maps and structural accessibility. (E) The optimal target (red arrow) was designated, after which (F) participants were randomized, with allocation concealed, to receive aiTBS or sham stimulation.

### 2.7. Outcomes

The primary outcome was change in Alzheimer Disease Assessment Scale–Cognitive Subscale (ADAS-Cog) after the 15-day treatment. Secondary outcomes included the change in ADAS-Cog score at the 3-month follow-up, as well as changes in MMSE at both the post-treatment and 3-month follow-up. We also assessed changes on a range of other measures, including the Montreal Cognitive Assessment (MoCA), Neuropsychiatric Inventory (NPI), Auditory Verbal Learning Test (AVLT), Trail Making Test (TMT), and Digit Span Test (DST). All adverse events happened during the intervention and within 3-month follow-up were recorded for evaluation of the safety of this protocol.

### 2.8. Statistical Analysis

This study did not include a formal hypothesis-testing sample size calculation. A total of 45 participants (≈15 per group) was selected pragmatically to evaluate the change of 15-day ADAS-Cog, and obtain preliminary effect-size estimates to inform the design of subsequent trials.

Efficacy analyses used a modified intention-to-treat population, excluding individuals who (1) did not receive any treatment after randomization or (2) were deemed ineligible post-randomization. Baseline characteristics were compared using ANOVA for continuous variables and chi-square (χ²) or Fisher’s exact tests for categorical variables. Primary and secondary outcomes were analyzed using linear mixed models, with Group (DMN, FPN, Sham), Time (baseline, 15-day, 3-month) and Group × Time interaction as fixed effect, participants as random effect. Response rates were compared with χ² tests. The significance level was 5% (two-tailed) with 95% confidence intervals (CIs). Pairwise comparisons were Bonferroni corrected. Statistical analyses were performed using SAS, version 9.4.

## 3. Results

### 3.1 Recruitment and Baseline Information

During the enrollment period from September 2023 to November 2024, 87 participants were screened and 46 participants were enrolled and randomized to receive active DMN (n = 15), active FPN (n = 17), or sham (n = 14) stimulation (Figure 2). Following randomization, one participant in the DMN group withdrew consent prior to receiving any intervention. Additionally, one participant was retrospectively found to have violated a key eligibility criterion and was consequently excluded from the modified intention-to-treat (mITT) analysis. The remaining 44 participants all completed the full treatment program. Baseline demographic and clinical characteristics were well-balanced across the three groups, as summarized in Table 1.

**Figure 2.**
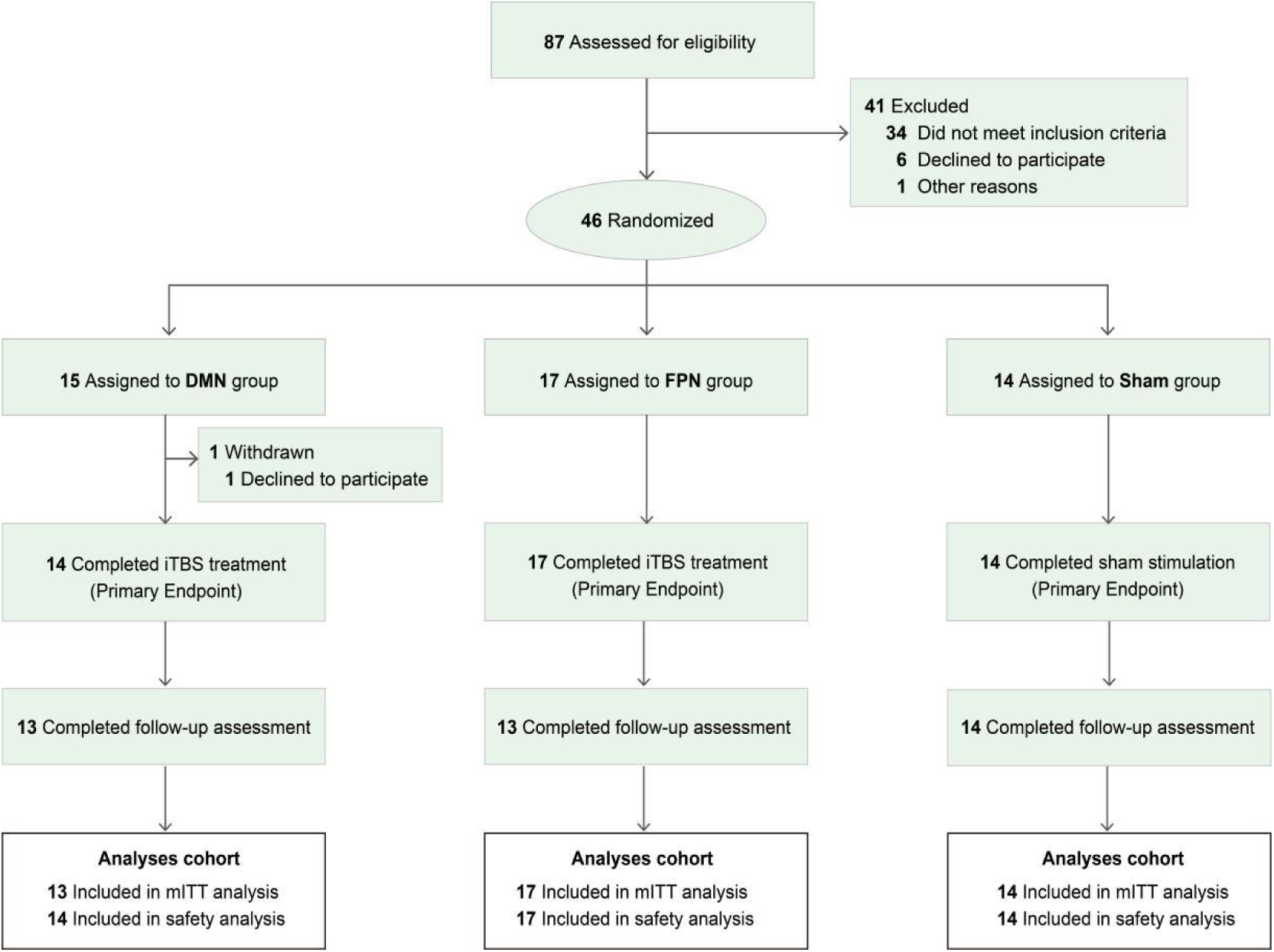
CONSORT trial flow diagram.

**Table 1.** Demographic and clinical characteristics at baseline.

| Measures | DMN, mean (SD)<br>(n = 14) | FPN, mean (SD)<br>(n = 17) | Sham, mean (SD)<br>(n = 14) | P value |
| --- | --- | --- | --- | --- |
| Age (years) | 69.15 (8.10) | 69.76 (7.84) | 66.21 (7.82) | 0.422 |
| Education (years) | 14.00 (2.68) | 13.82 (3.47) | 14.00 (2.15) | 0.981 |
| Gender (male/female) | 2/11 | 6/11 | 4/10 | 0.475 |
| ADAS-Cog | 17.15 (3.85) | 19.59 (4.66) | 17.43 (3.59) | 0.207 |
| MMSE | 22.46 (1.81) | 21.71 (1.90) | 20.93 (1.33) | 0.078 |
| MoCA | 17.92 (2.78) | 18.00 (2.72) | 18.00 (2.69) | 0.996 |
| NPI | 0.54 (1.33) | 1.24 (2.14) | 0.93 (2.30) | 0.641 |
| AVLT |  |  |  |  |
| -Immediate | 14.15 (2.70) | 13.24 (3.72) | 14.21 (3.36) | 0.657 |
| -Delay | 0.31 (0.86) | 0.24 (0.56) | 0.21 (0.58) | 0.930 |
| -Recognition | 2.31 (2.78) | 3.29 (2.80) | 3.21 (2.97) | 0.603 |
| Digital span test |  |  |  |  |
| -Forward | 7.92 (1.12) | 7.76 (0.97) | 8.07 (1.00) | 0.709 |
| -Backward | 4.08 (1.19) | 4.71 (0.99) | 4.07 (1.00) | 0.165 |
| TMT-A | 66.69 (23.75) | 69.00 (24.16) | 79.36 (20.46) | 0.309 |
| TMT-B | 137.31 (74.09) | 160.29 (83.49) | 158.21 (83.43) | 0.711 |

### 3.2 Treatment Efficacy

As a primary outcome, significant improvements on the ADAS-Cog scores at 15-day post-treatment were observed for both the DMN (estimated difference = -2.85, *p* < 0.0001, 95% CI = -4.13- -1.57) and FPN groups (estimated difference = -2.41, *p* < 0.0001, 95% CI = -3.53- -1.29), but not for the sham group (estimated difference = 0.21, *p* = 0.7301, 95% CI = -1.01 - 1.45) (Figure 3, Table 2). Notably, in the between-group analyses, both the DMN and FPN groups showed significantly greater improvement than the sham group (DMN vs Sham: estimated difference = -3.06, *p* = 0.0020, 95 % CI = -4.84 - -1.28; FPN vs Sham: estimated difference = -2.63, *p* = 0.0048, 95 % CI = -4.29- -0.96).

**Figure 3.**
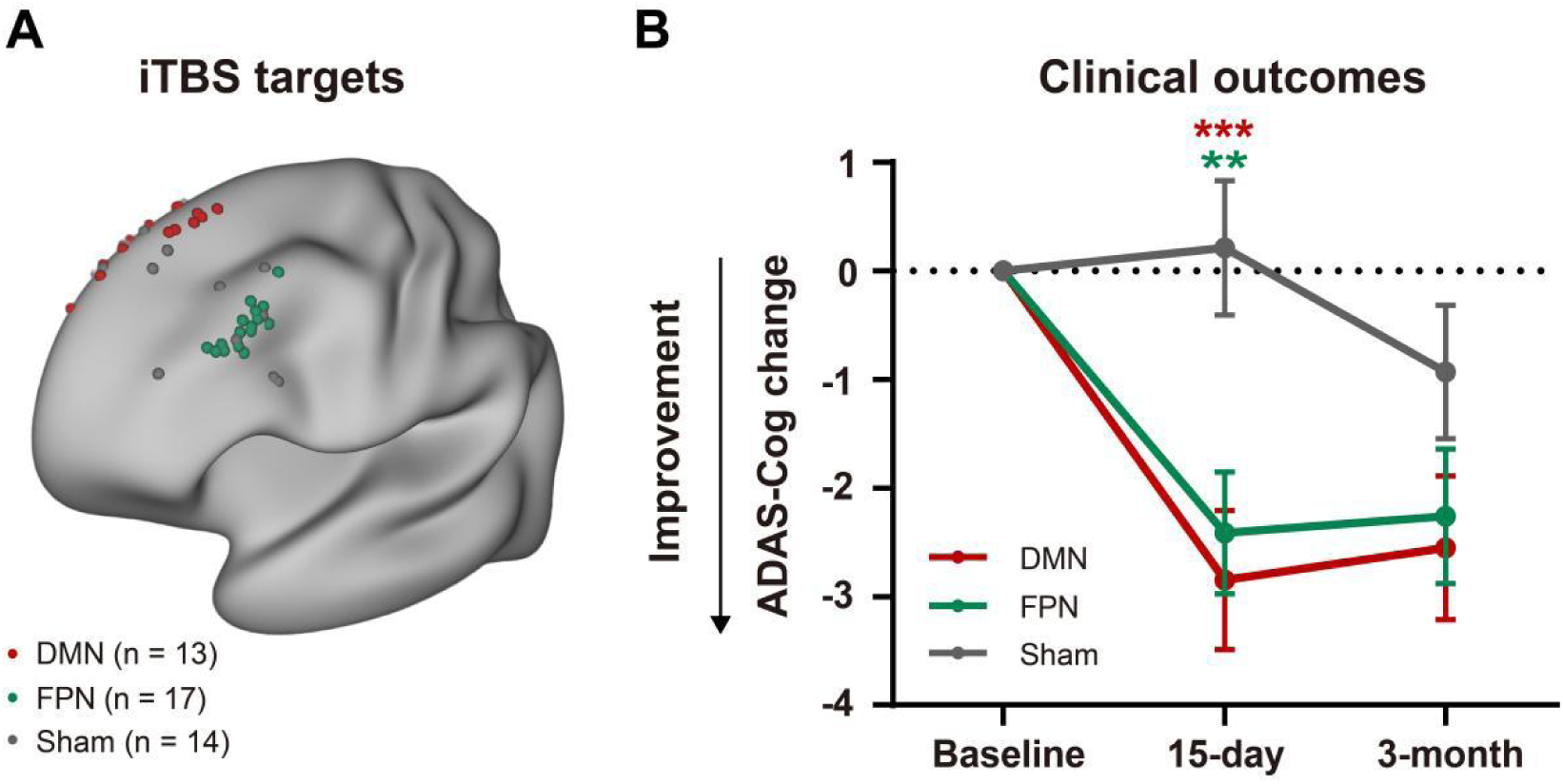
aiTBS targeting personalized DMN or FPN nodes improves ADAS-Cog scores in mild AD. A. Personalized stimulation targets within the default mode network (DMN; red, n = 13), fronto-parietal network (FPN; green, n = 17) and sham stimulation targets (grey, n = 14) were displayed on a standard left hemisphere surface. B. Mean score changes in ADAS-Cog at post-treatment and 3-month follow-up for the DMN (red), FPN (green) and sham (grey) groups. Negative values indicate improvement; error bars represent the standard error.

**Table 2.**
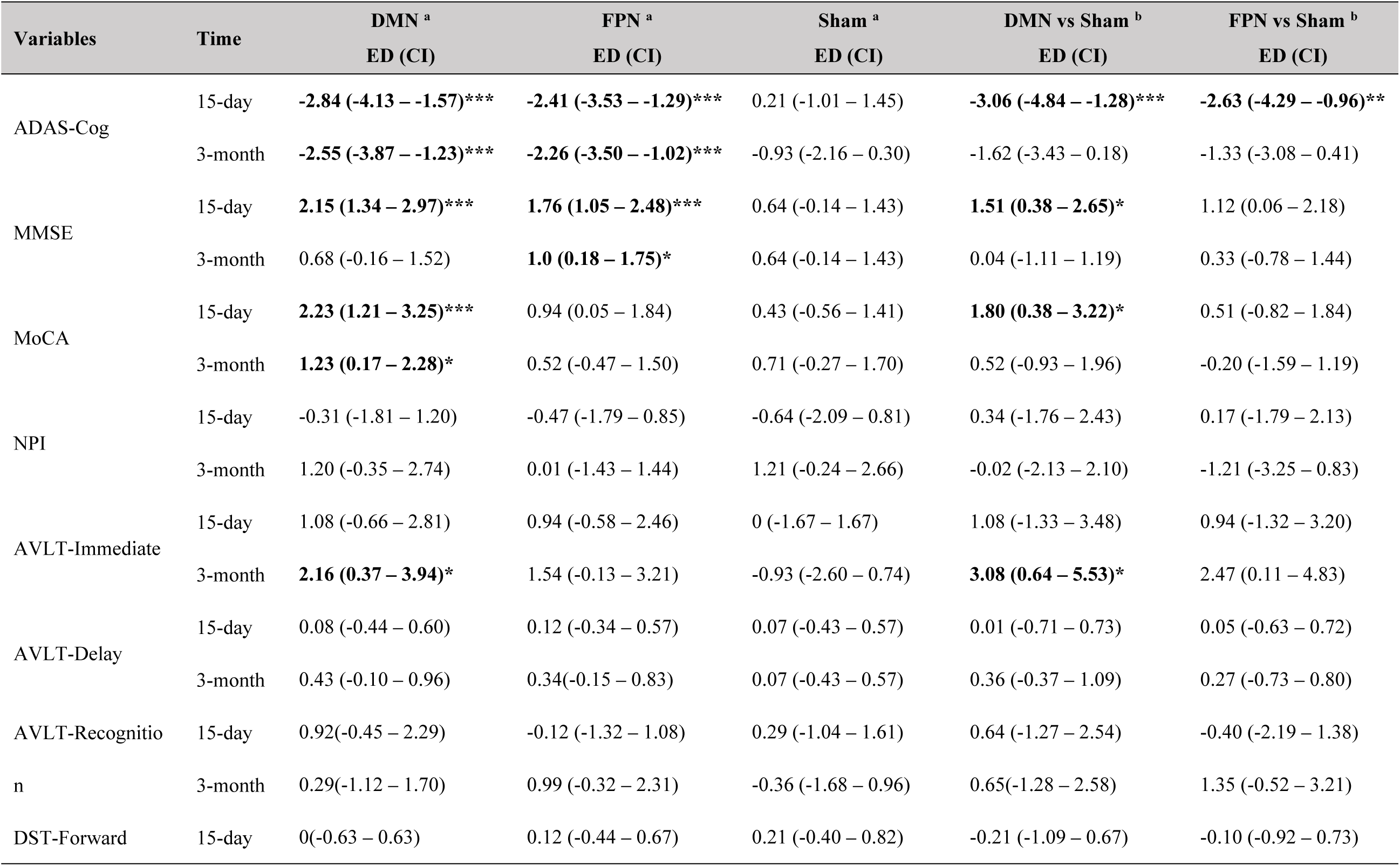
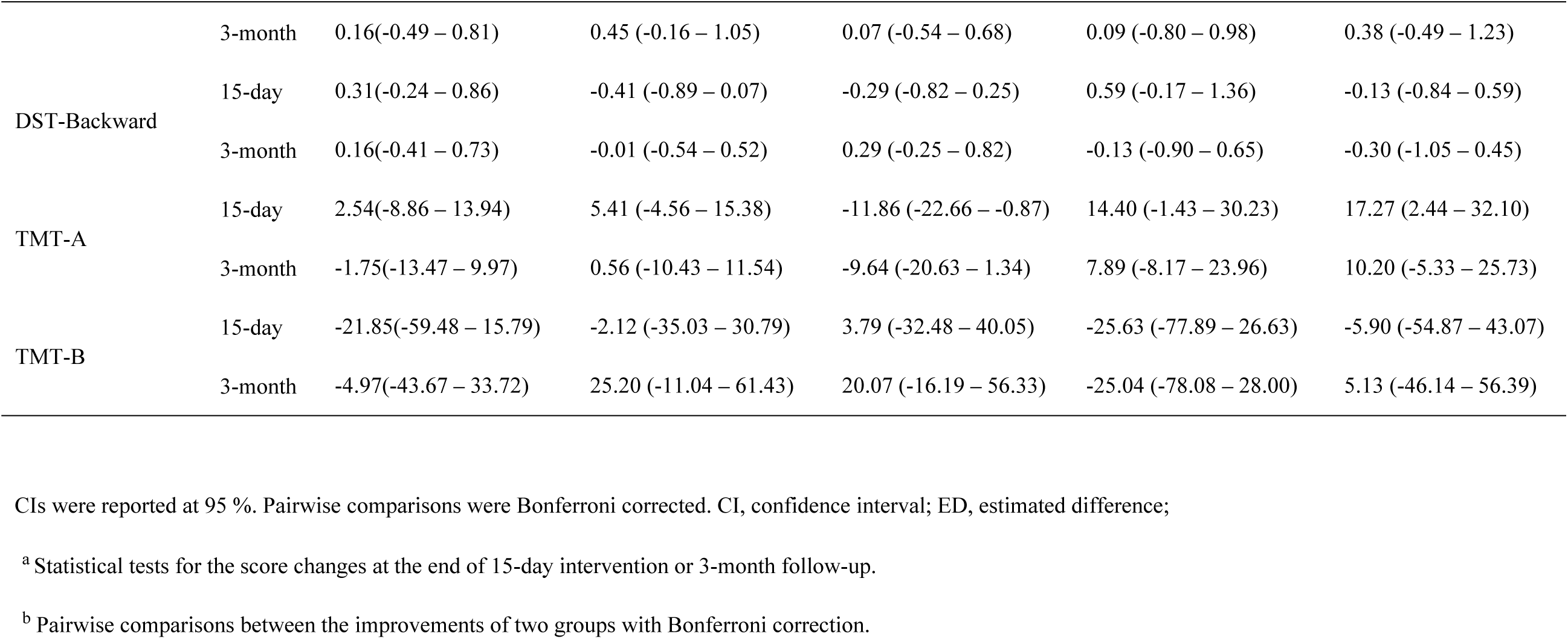
Effects of personalized aiTBS in each measurement for all groups.

Analysis of the secondary outcome, ADAS-Cog scores at the 3-month follow-up, revealed sustained cognitive improvements within both the DMN and FPN active stimulation groups. Significant within-group improvements from baseline were maintained in the DMN group (estimated difference = -2.55, *p* = 0.0002, 95 % CI = -3.87- -1.23) and the FPN group (estimated difference = -2.26, *p* = 0.0005, 95 % CI = -3.50 - -1.02), whereas the sham group showed no significant change (estimated difference = -0.93, *p* = 0.1376, 95 % CI = -2.16 - 0.30). However, in the between-group comparisons, the magnitude of improvement in the DMN and FPN groups was not statistically superior to that observed in the sham control at this 3-month time point (DMN vs Sham: estimated difference = -1.33, p = 0.1548, 95 % CI = -3.08- 0.41; FPN vs Sham: estimated difference = -1.62, *p* = 0.2650, 95 % CI = -3.43- 0.18).

Analysis of MMSE scores, a secondary outcome, provided further evidence of cognitive improvement. Significant within-group increases were observed in the DMN group immediately post-treatment (estimated difference = 2.15, *p* < 0.0001, 95 % CI = 1.34 - 2.97), and in the FPN group at both the post-treatment (estimated difference = 1.76, *p* < 0.0001, 95 % CI = 1.05 - 2.48) and 3-month follow-up (estimated difference = 1.0, *p* = 0.0166, 95 % CI = 0.18- 1.75). In contrast, the sham group showed no significant within-group improvement at either time point (post-treatment: estimated difference = 0.64, *p* = 0.1080, 95 % CI = -0.14 - 1.43; 3-month: estimated difference = 0.64, *p* = 0.1080, 95 % CI = -0.14 - 1.43). Between-group comparisons confirmed that the improvement in the DMN group was significantly greater than that in the sham group at post-treatment (estimated difference = 1.51, *p* = 0.0194, 95 % CI = 0.38 - 2.65). No other between-group comparisons reached statistical significance.

On the other secondary outcomes, the MoCA showed a significant improvement for the DMN group relative to sham at post-treatment (estimated difference = 1.80, *p* = 0.0272, 95 % CI = 0.38 - 3.22), but not at the 3-month follow-up (estimated difference = 0.52, *p* = 0.9580, 95 % CI = -0.93- 1.96). For the AVLT-Immediate, the DMN group demonstrated significant superiority over sham at the 3-month follow-up (estimated difference = 3.08, *p* = 0.0280, 95 % CI = 0.64 - 5.53) but not at the post-treatment (estimated difference = 2.47, *p* = 0.7518, 95 % CI = 0.11- 4.83). No statistically significant between-group differences were found at either time point for the AVLT-Delay, AVLT-Recognition, NPI, Trail TMT, or DST.

Responder analysis further substantiated the treatment efficacy (Figure 4). A clinically meaningful improvement, defined as a reduction of ≥3 points on the ADAS-Cog, was achieved by a markedly higher proportion of participants in the active groups at the 15-day post-treatment: 38.46% in the DMN group and 47.06% in the FPN group, compared to 0% in the sham group. Notably, all participants (100%) in the DMN group and 70.59% in the FPN group demonstrated an improvement of at least 2 points on the ADAS-Cog, compared to only 7.14% in the sham group.

**Figure 4.**
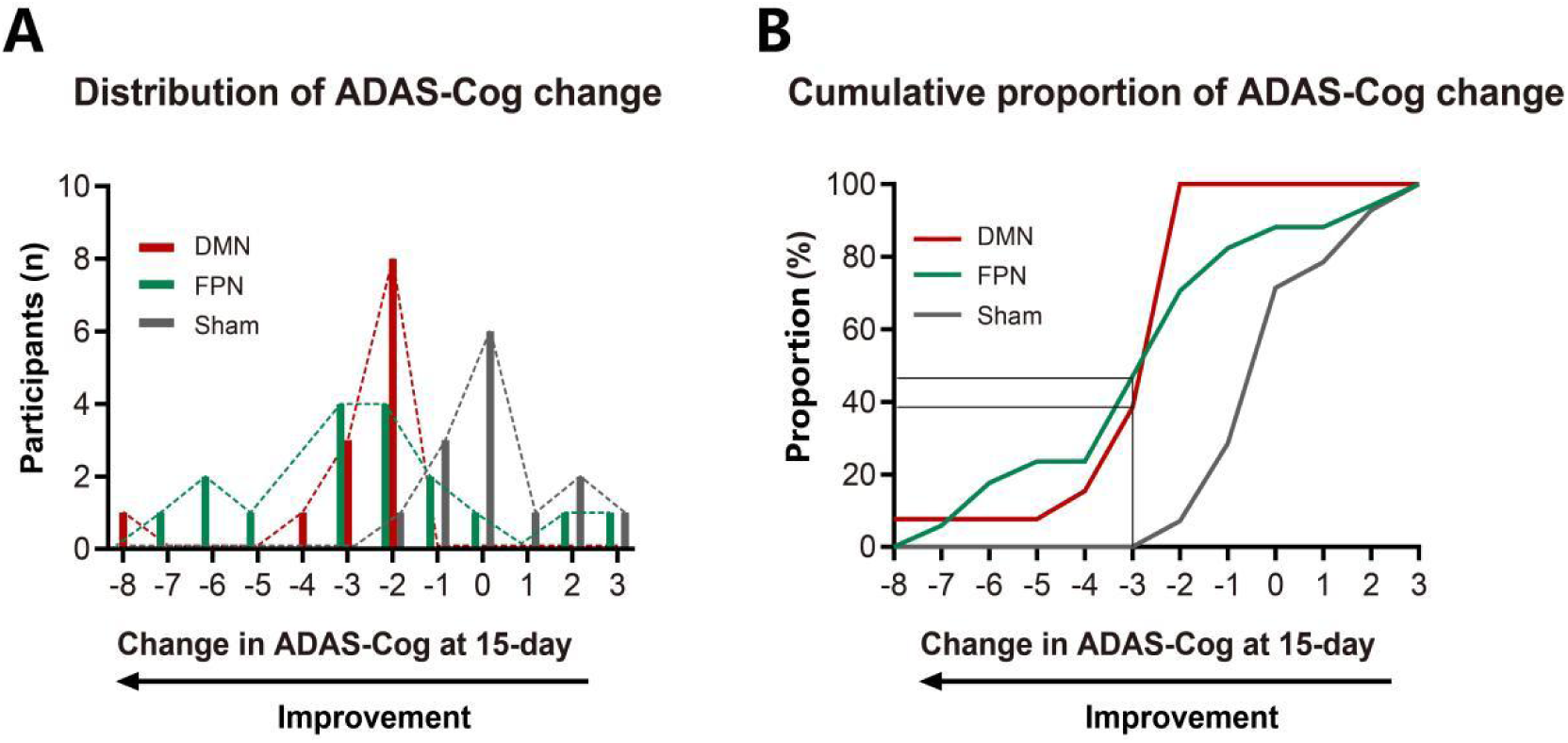
Responder analysis of ADAS-Cog changes at post-treatment. A. Distribution of individual ADAS-Cog score changes at post-treatment for the DMN (red), FPN (green) and sham (grey) groups. Negative values indicate cognitive improvement. B. Cumulative proportion of participants achieving at least the indicated improvement in ADAS-Cog score at post-treatment. Both active treatment groups (DMN and FPN) display a left-shifted distribution and markedly higher cumulative incidence of improvement compared with sham group.

### 3.3 Safety

Safety was evaluated in all 45 participants who received at least one session of stimulation. One serious adverse event (a postoperative inguinal hernia) occurred during follow-up and was judged unrelated to the intervention; it resolved fully after surgical treatment. The most common adverse event was transient scalp discomfort, reported in 13% (6/45) of participants. Isolated, self-resolving cases of dizziness and fatigue were also recorded, with some assessed as unrelated to stimulation. Though a greater number of adverse events were reported in the FPN group (n = 6) compared to the DMN (n = 2) and sham (n = 1) groups, this distribution was not statistically significant (Fisher’s exact test, *p* = 0.146). The overall safety profile supports the feasibility of delivering 7200 pulses of daily aiTBS to either the left DMPFC or DLPFC in patients with mild AD.

## 4. Discussion

In this randomized, sham-controlled, three-arm clinical trial, a 15-day aiTBS targeting the personalized DMN or FPN nodes in patients with mild AD yielded significantly greater cognitive improvement compared with sham stimulation and was well-tolerated. These findings support the promising efficacy and safety of this personalized, network-based, aiTBS as a therapeutic intervention for AD, and as anti-amyloid therapies enter real-world practice with acceptable short-term safety, circuit-based neuromodulation may offer a complementary approach to address persistent cognitive symptoms.^34^

This trial was designed to target specific functional networks implicated in the cognitive impairment of AD. Both the DMN and FPN active aiTBS groups demonstrated significantly greater improvement on the primary outcome (change in ADAS-Cog score) than the sham group. The DMN group showed a −3.06-point advantage over sham (95% CI, −4.84 to −1.28), and the FPN group a −2.63-point advantage (95% CI, −4.29 to −0.96). Despite the short treatment duration in this trial, the magnitude of effect approximated the pooled mean difference reported in a meta-analysis of rTMS combined with cognitive training delivered for ≥4 weeks (−2.49 points; 95% CI, −3.77 to −1.22).^35^ Responder analyses revealed clinically meaningful improvement (benefit of ≤ -3 point on ADAS-Cog) in 38.46% of the DMN group and 47.06% of the FPN group, similar to that observed after 6 weeks of multi-site 10-Hz rTMS in patients with milder AD (baseline ADAS-Cog ≤30).^36^

The cognitive improvements resulting from DMN or FPN stimulation are likely mediated by neuroplastic changes within these networks, a mechanism consistent with the known pathophysiology of AD.^22^ The DMN, essential for memory storage and consolidation, is linked to AD pathogenesis when disrupted.^37^ Decreased DMN connectivity in AD has been consistently demonstrated, especially between its prefrontal and posterior hubs.^4,38,39^ While previous rTMS studies have targeted posterior DMN nodes such as the precuneus^14,40,41^, parietal cortex^15,42^, and angular gyrus^43^, here we examined a novel prefrontal DMN target, selected for its strong functional connectivity with key posterior DMN hubs. Significant improvements were observed across ADAS-Cog, MMSE, and MoCA following DMPFC stimulation, suggesting that prefrontal node targeting can also effectively modulate DMN function and yield meaningful clinical benefits. Together with evidence from posterior DMN stimulation, these findings suggest DMN as a promising therapeutic target for AD.

The FPN, essential for executive functions and working memory, is impaired in AD.^44^ Clinical studies, including randomized trials, indicate that rTMS applied to the DLPFC within the PFN (FPN) is associated with improvements in global cognition^16,17^, memory^31,32,45^, language^46^, sleep quality^47^, and apathy.^48^ Yet results have been mixed, probably due to the large variability in stimulation targets. To enhance intended circuit engagement, we used precision functional mapping technique to localize each participant’s FPN node. The 15-day aiTBS treatment targeting the FPN produced a significant improvement compared to the sham group on ADAS-Cog but not on MMSE or MoCA, which may reflect differences in the sensitivity^49^ of cognitive outcome measures over short intervals. A meta-analysis showed that, relative to sham stimulation, high-frequency rTMS improved ADAS-cog but not MMSE.^50^

The efficacy of TMS is highly dependent on protocol optimization^12^, yet a lack of standardized parameters persists across targets, duration, and pulse number.^18, 20^ Our study introduces an aiTBS protocol designed to overcome this limitation. By integrating precision targeting with a high daily dose (7,200 pulses) over a condensed 15-day course, this trial achieved significant cognitive improvement. The sham group showed no significant change in cognitive scores despite stable AD medications, suggesting that observed gains were not attributable to concomitant therapy. Mechanistically, aiTBS has been shown to induce resting-state connectivity reconfiguration that tracks cognitive improvement in amnestic MCI, supporting circuit engagement as a plausible mediator of benefit.^51^ Although 15 days of high-dose aiTBS improved cognition in mild AD, between-group differences were no longer significant at 3-month follow-up, indicating that maintenance strategies may be required to sustain benefit and should be evaluated in future studies.

Despite the promising results, this study has several limitations. First, the generalizability of the findings is constrained by the relatively small sample size and the single-center design. Large-scale, multi-center trials are needed to further confirm the efficacy of this approach. Second, while the clinical diagnosis of AD was supported by amyloid PET or CSF biomarkers, the use of different biomarkers may introduce potential heterogeneity into the study population.

In conclusion, this single-center, randomized, sham-controlled clinical trial demonstrates that a 15-day high-dose aiTBS targeting the personalized DMN or FPN nodes is an effective and safe intervention for cognitive enhancement in mild AD. These findings suggest the DMN and FPN networks as the promising neuromodulation targets for AD.

## Acknowledgements

The authors thank all the study participants and their families.

## Consent Statement

All human subjects provided informed consent。

## Data Availability

The data that support the main findings of this study are available from the corresponding author, on reasonable request.

## References

1. Seeley WW, Crawford RK, Zhou J, Miller BL, Greicius MD. Neurodegenerative diseases target large-scale human brain networks. Neuron. 2009;62:42–52. doi:10.1016/j.neuron.2009.03.024.

2. Palmqvist S, Scholl M, Strandberg O, et al. Earliest accumulation of beta-amyloid occurs within the default-mode network and concurrently affects brain connectivity. Nat Commun. 2017;8:1214. doi:10.1038/s41467-017-01150-x.

3. Jagust W. Imaging the evolution and pathophysiology of Alzheimer disease. Nat Rev Neurosci. 2018;19:687–700. doi:10.1038/s41583-018-0067-3.

4. Greicius MD, Srivastava G, Reiss AL, Menon V. Default-mode network activity distinguishes Alzheimer’s disease from healthy aging: evidence from functional MRI. Proc Natl Acad Sci U S A. 2004;101:4637–4642. doi:10.1073/pnas.0308627101.

5. Tetreault AM, Phan T, Orlando D, et al. Network localization of clinical, cognitive, and neuropsychiatric symptoms in Alzheimer’s disease. Brain. 2020;143:1249–1260. doi:10.1093/brain/awaa058.

6. Buckner RL, Sepulcre J, Talukdar T, et al. Cortical hubs revealed by intrinsic functional connectivity: mapping, assessment of stability, and relation to Alzheimer’s disease. J Neurosci. 2009;29:1860–1873. doi:10.1523/JNEUROSCI.5062-08.2009.

7. Vogel JW, Iturria-Medina Y, Strandberg OT, et al. Spread of pathological tau proteins through communicating neurons in human Alzheimer’s disease. Nat Commun. 2020;11:2612. doi:10.1038/s41467-020-15701-2.

8. Jones DT, Knopman DS, Gunter JL, et al. Cascading network failure across the Alzheimer’s disease spectrum. Brain. 2016;139:547–562. doi:10.1093/brain/awv338.

9. Jobson DD, Hase Y, Clarkson AN, Kalaria RN. The role of the medial prefrontal cortex in cognition, ageing and dementia. Brain Commun. 2021;3:fcab125. doi: 10.1093/braincomms/fcab125.

10. Friedman NP, Robbins TW. The role of prefrontal cortex in cognitive control and executive function. Neuropsychopharmacology. 2022;47:72–89. doi:10.1038/s41386-021-01132-0.

11. Rossi S, Antal A, Bestmann S, et al. Safety and recommendations for TMS use in healthy subjects and patient populations, with updates on training, ethical and regulatory issues: Expert Guidelines. Clin Neurophysiol. 2021;132:269–306. doi:10.1016/j.clinph.2020.10.003.

12. Lefaucheur JP, Aleman A, Baeken C, et al. Evidence-based guidelines on the therapeutic use of repetitive transcranial magnetic stimulation (rTMS): An update (2014-2018). Clin Neurophysiol. 2020;131:474–528. doi:10.1016/j.clinph.2019.11.002.

13. Menardi A, Rossi S, Koch G, et al. Toward noninvasive brain stimulation 2.0 in Alzheimer’s disease. Ageing Res Rev. 2022;75:101555. doi:10.1016/j.arr.2021.101555.

14. Koch G, Casula EP, Bonni S, et al. Precuneus magnetic stimulation for Alzheimer’s disease: A randomized, sham-controlled trial. Brain. 2022;145:3776–3786. doi:10.1093/brain/awac285.

15. Jung YH, Jang H, Park S, et al. Effectiveness of personalized hippocampal network-targeted stimulation in Alzheimer disease: A randomized clinical trial. JAMA Netw Open. 2024;7:e249220. doi:10.1001/jamanetworkopen.2024.9220.

16. Li X, Qi G, Yu C, et al. Cortical plasticity is correlated with cognitive improvement in Alzheimer’s disease patients after rTMS treatment. Brain Stimul. 2021;14:503–510. doi:10.1016/j.brs.2021.01.012.

17. Wu X, Yan Y, Hu P, et al. Effects of a periodic intermittent theta burst stimulation in Alzheimer’s disease. Gen Psychiatr. 2024;37:e101106. doi:10.1136/gpsych-2023-101106.

18. Zhang Y, Dong K, Yang J, et al. Comparative efficacy of rTMS on different targets in Alzheimer’s disease: A systematic review and meta-analysis. Front Aging Neurosci. 2025;17:1536573. doi:10.3389/fnagi.2025.1536573.

19. Menardi A, Dotti L, Ambrosini E, Vallesi A. Transcranial magnetic stimulation treatment in Alzheimer’s disease: A meta-analysis of its efficacy as a function of protocol characteristics and degree of personalization. J Neurol. 2022;269:5283–5301. doi:10.1007/s00415-022-11236-2.

20. Li S, Lan X, Liu Y, et al. Unlocking the potential of repetitive transcranial magnetic stimulation in Alzheimer’s disease: A meta-analysis of randomized clinical trials to optimize intervention strategies. J Alzheimers Dis. 2024;98:481–503. doi:10.3233/JAD-231031.

21. Rektorova I, Pupikova M, Fleury L, Brabenec L, Hummel FC. Non-invasive brain stimulation: current and future applications in neurology. Nat Rev Neurol. 2025;21:669–686. doi:10.1038/s41582-025-01137-z.

22. Eldaief MC, McMains S, Izquierdo-Garcia D, Daneshzand M, Nummenmaa A, Braga RM. Network-specific metabolic and haemodynamic effects elicited by non-invasive brain stimulation. Nat Ment Health. 2023;1:346–360. doi:10.1038/s44220-023-00046-8.

23. Wang D, Buckner RL, Fox MD, et al. Parcellating cortical functional networks in individuals. Nat Neurosci. 2015;18:1853–1860. doi:10.1038/nn.4164.

24. Gordon EM, Laumann TO, Gilmore AW, et al. Precision functional mapping of individual human brains. Neuron. 2017;95:791–807.e7. doi:10.1016/j.neuron.2017.07.011.

25. Mir-Moghtadaei A, Caballero R, Fried P, et al. Concordance between beamF3 and MRI-neuronavigated target sites for repetitive transcranial magnetic stimulation of the left dorsolateral prefrontal cortex. Brain Stimul. 2015;8:965–973. doi:10.1016/j.brs.2015.05.008.

26. Cash RFH, Weigand A, Zalesky A, et al. Using brain imaging to improve spatial targeting of transcranial magnetic stimulation for depression. Biol Psychiatry. 2021;90:689–700. doi:10.1016/j.biopsych.2020.05.033.

27. Moussavi Z, Uehara M, Rutherford G, et al. Repetitive transcranial magnetic stimulation as a treatment for Alzheimer’s disease: A randomized placebo-controlled double-blind clinical trial. Neurotherapeutics. 2024;21:e00331. doi:10.1016/j.neurot.2024.e00331.

28. Saitoh Y, Hosomi K, Mano T, et al. Randomized, sham-controlled, clinical trial of repetitive transcranial magnetic stimulation for patients with Alzheimer’s dementia in Japan. Front Aging Neurosci. 2022;14:993306. doi:10.3389/fnagi.2022.993306.

29. Caulfield KA, Fleischmann HH, George MS, McTeague LM. A transdiagnostic review of safety, efficacy, and parameter space in accelerated transcranial magnetic stimulation. J Psychiatr Res. 2022;152:384–396. doi:10.1016/j.jpsychires.2022.06.038.

30. Cole EJ, Phillips AL, Bentzley BS, et al. Stanford neuromodulation therapy (SNT): A double-blind randomized controlled trial. Am J Psychiatry. 2022;179:132–141. doi:10.1176/appi.ajp.2021.20101429.

31. Lin H, Liang J, Wang Q, et al. Effects of accelerated intermittent theta-burst stimulation in modulating brain of Alzheimer’s disease. Cereb Cortex. 2024;34:bhae106. doi:10.1093/cercor/bhae106.

32. Wu X, Ji GJ, Geng Z, et al. Accelerated intermittent theta-burst stimulation broadly ameliorates symptoms and cognition in Alzheimer’s disease: A randomized controlled trial. Brain Stimul. 2022;15:35–45. doi:10.1016/j.brs.2021.11.007.

33. Yeo BT, Krienen FM, Sepulcre J, et al. The organization of the human cerebral cortex estimated by intrinsic functional connectivity. J Neurophysiol. 2011;106:1125–1165. doi:10.1152/jn.00338.2011.

34. Li LL, Wang RZ, Wang Z, et al. Safety and short-term outcomes of lecanemab for Alzheimer’s disease in China: a multicentre study. Brain. 2025;awaf427. doi:10.1093/brain/awaf427.

35. Liu G, Xue B, Guan Y, Luo X. Effects of repetitive transcranial magnetic stimulation combined with cognitive training on cognitive function in patients with Alzheimer’s disease: A systematic review and meta-analysis. Front Aging Neurosci. 2023;15:1254523. doi:10.3389/fnagi.2023.1254523.

36. Sabbagh M, Sadowsky C, Tousi B, et al. Effects of a combined transcranial magnetic stimulation (TMS) and cognitive training intervention in patients with Alzheimer’s disease. Alzheimers Dement. 2020;16:641–650. doi:10.1016/j.jalz.2019.08.197.

37. Dennis EL, Thompson PM. Functional brain connectivity using fMRI in aging and Alzheimer’s disease. Neuropsychol Rev. 2014;24:49–62. doi:10.1007/s11065-014-9249-6.

38. Wang L, Zang Y, He Y, et al. Changes in hippocampal connectivity in the early stages of Alzheimer’s disease: evidence from resting state fMRI. Neuroimage. 2006;31:496–504. doi:10.1016/j.neuroimage.2005.12.033.

39. Cai Y, Yang J, Liang L, et al. White matter hyperintensities correlate with accelerated tau accumulation in posterior cortical regions in Alzheimer’s disease. Alzheimers Dement. 2026;21(Suppl 2):e104870. doi:10.1002/alz70856_104870.

40. Koch G, Bonni S, Pellicciari MC, et al. Transcranial magnetic stimulation of the precuneus enhances memory and neural activity in prodromal Alzheimer’s disease. Neuroimage. 2018;169:302–311. doi:10.1016/j.neuroimage.2017.12.048.

41. Koch G, Casula EP, Bonni S, et al. Effects of 52 weeks of precuneus rTMS in Alzheimer’s disease patients: A randomized trial. Alzheimers Res Ther. 2025;17:69. doi:10.1186/s13195-025-01709-7.

42. Jia Y, Xu L, Yang K, et al. Precision repetitive transcranial magnetic stimulation over the left parietal cortex improves memory in Alzheimer’s disease: A randomized, double-blind, sham-controlled study. Front Aging Neurosci. 2021;13:693611. doi:10.3389/fnagi.2021.693611.

43. Liu C, Han T, Xu Z, et al. Modulating gamma oscillations promotes brain connectivity to improve cognitive impairment. Cereb Cortex. 2022;32:2644–2656. doi:10.1093/cercor/bhab371.

44. Dosenbach NU, Fair DA, Miezin FM, et al. Distinct brain networks for adaptive and stable task control in humans. Proc Natl Acad Sci U S A. 2007;104:11073–11078. doi:10.1073/pnas.0704320104.

45. Bagattini C, Zanni M, Barocco F, et al. Enhancing cognitive training effects in Alzheimer’s disease: rTMS as an add-on treatment. Brain Stimul. 2020;13:1655–1664. doi:10.1016/j.brs.2020.09.010.

46. Cotelli M, Manenti R, Cappa SF, et al. Effect of transcranial magnetic stimulation on action naming in patients with Alzheimer disease. Arch Neurol. 2006;63:1602–1604. doi:10.1001/archneur.63.11.1602.

47. Zhou X, Wang Y, Lv S, et al. Transcranial magnetic stimulation for sleep disorders in Alzheimer’s disease: A double-blind, randomized, and sham-controlled pilot study. Neurosci Lett. 2022;766:136337. doi:10.1016/j.neulet.2021.136337.

48. Padala PR, Boozer EM, Lensing SY, et al. Neuromodulation for apathy in Alzheimer’s disease: A double-blind, randomized, sham-controlled pilot study. J Alzheimers Dis. 2020;77:1483–1493. doi:10.3233/JAD-200640.

49. Levine SZ, Yoshida K, Goldberg Y, et al. Linking the Mini-Mental State Examination, the Alzheimer’s Disease Assessment Scale-Cognitive Subscale and the Severe Impairment Battery: evidence from individual participant data from five randomised clinical trials of donepezil. Evid Based Ment Health. 2021;24:56–61. doi:10.1136/ebmental-2020-300184.

50. Dong X, Yan L, Huang L, et al. Repetitive transcranial magnetic stimulation for the treatment of Alzheimer’s disease: A systematic review and meta-analysis of randomized controlled trials. PLoS One. 2018;13:e0205704. doi:10.1371/journal.pone.0205704.

51. Aghamoosa S, Nolin SA, Chen AA, et al. Accelerated iTBS-induced changes in resting-state functional connectivity correspond with cognitive improvement in amnestic MCI. Brain Stimul. 2025;18:957–964. doi:10.1016/j.brs.2025.04.012.

